# High false-positive rate of fluorodeoxyglucose positron emission tomography to detect lymph node metastasis in microinvasive cervical cancer after conization

**DOI:** 10.1101/2021.02.19.21251455

**Authors:** Kidong Kim, Hayeon Kim, Hwajung Lee, Keun Ho Lee, Banghyun Lee, ChelHun Choi, Seung-Hyuk Shim, Jong-Min Lee, Miseon Kim, Jung-Yun Lee

## Abstract

We aimed to estimate the positive predictive value of fluorodeoxyglucose positron emission tomography (FDG PET) to detect lymph node metastasis in patients with microinvasive cervical cancer who recently underwent conization. We retrospectively collected data of patients fulfilling the following criteria: 1) cervical cancer stage 1A1 (by FIGO staging revised in 1994) without invasion of the lymphovascular space diagnosed by conization from September 2008 to July 2018, 2) FDG PET within 3 months after diagnosis, 3) lymph node metastasis suspected by FDG PET, and 4) histologic confirmation or follow-up imaging study for suspected lymph node metastasis. Lymph node metastasis was suspected in 31 regions in 18 patients; however, no true metastasis was found. In conclusion, lymph node metastasis suspected by FDG PET in microinvasive cervical cancer after conization might be false positive. Lymph node dissection should not be performed for such patients.

## Introduction

Patients with microinvasive cervical cancer without lymphovascular space invasion have a very low risk of lymph node metastasis and do not need to undergo lymph node dissection [1]. Rarely, preoperative fluorodeoxyglucose positron emission tomography (FDG PET) reveals suspected lymph node metastasis in patients with microinvasive cervical cancer. For these patients, it is unclear whether suspected lymph nodes should be resected. Omission of lymph node resection could result in bad prognosis for true metastasis, but lymph node resection for false-positive cases could increase surgery time, risk of organ injury, and lymphedema. The objective of this study was to estimate the positive predictive value of FDG PET to detect lymph node metastasis in patients with microinvasive cervical cancer.

## Methods

After the protocol was approved by the Institutional Review Board of each participating institute, we selected the patients satisfying the following eligibility criteria: 1) cervical cancer stage 1A1 (by FIGO staging revised in 1994) without lymphovascular space invasion diagnosed by conization from September 2008 to July 2018, 2) FDG PET within 3 months after diagnosis, 3) lymph node metastasis suspected by FDG PET, 4) Excision or follow-up imaging study for suspected lymph nodes. Suspected lymph node metastasis was defined as moderately to markedly increased FDG uptake in the lymph node area, excluding physiologic uptake in bowel or urinary tract. Acquisition of informed consent was waived by the Institutional Review Board.

We collected data, including age, menopausal status, tumor width, invasion depth, margin status of conization specimen, result of FDG PET, and result of surgical resection or follow-up imaging study. We summarized patient characteristics and estimated the positive predictive value of FDG PET using the result of surgical resection or follow-up imaging study as a reference.

We classified lymph nodes into 3 regions: left pelvic, right pelvic, and para-aortic. For example, if a patient was suspected with lymph node metastasis in the right and left pelvic areas, 2 regions from 1 patient were included in the analysis. We examined the positive predictive value at the region and patient levels.

## Results

Eighteen patients from 6 institutes satisfied the eligibility criteria and were included in this study. The median age was 35 years, and most patients were premenopausal. The median tumor width and median invasion depth were both 1 mm. The resection margin of the conization specimen was involved by cervical intraepithelial neoplasia or cancer in about two-thirds of patients. Suspected lymph node metastasis was detected by FDG PET mainly in the pelvic area but rarely in the para-aortic area. The results of surgical resection were used as reference in most cases (Table 1). Suspected lymph node metastasis was present in 31 regions in 18 patients; however, no true metastasis was found.

**Table 1.** Patient characteristics (n = 18)

| Characteristics | Value |
| --- | --- |
| Age, y, median (range) | 34.5 (27 – 63) |
| Menopausal status, n (%) |  |
| Premenopause | 17 (94) |
| Postmenopause | 1 (6) |
| Tumor width, mm, median (range) | 1 (0.2 – 7) |
| Invasion depth, mm, median (range) | 1 (0.1 – 3) |
| Margin status of conization specimen, n (%) |  |
| Negative | 7 (39) |
| Cervical intraepithelial neoplasia | 7 (39) |
| Cancer | 4 (22) |
| Interval from conization to FDG PET, day, median (range) | 13.5 (1 – 90) |
| Location of suspected lymph node metastasis, n (%) |  |
| Left pelvic | 15 (83) |
| Right pelvic | 13 (72) |
| Para-aortic | 3 (17) |
| Method to confirm lymph node metastasis, n (%) |  |
| Surgical resection | 13 (72) |
| Follow-up imaging study | 5 (28) |
| FDG PET, fluorodeoxyglucose positron emission tomography |  |

## Discussion

Our data suggests that the detection of suspected lymph node metastasis by FDG PET in patients with microinvasive cervical cancer who recently underwent conization might be false positive. For such patients, we may omit surgical resection of suspected lymph nodes. Clinicians who are well experienced may perform lymph node sampling. However, we believe that lymph node dissection should not be performed for these patients considering the sequalae of the procedure.

We believe that false-positive FDG uptake in lymph nodes is due to conization. Like other excisional procedures, cervical conization induces locoregional inflammatory reaction, resulting in FDG uptake in regional lymph nodes [2]. Because most cases of microinvasive cervical cancer are diagnosed by conization, FDG PET scan as a preoperative work-up could falsely diagnose inflammatory FDG uptake as lymph node metastasis.

In conclusion, lymph node metastasis suspected by FDG PET in microinvasive cervical cancer after conization could be false positive. Hence, for these patients, lymph node dissection should not be performed.

## Data Availability

Data is available upon reasonable request to the corresponding author.

## Acknowledgement

None

## Ethical approval

The protocol was approved by the Institutional Review Board of each participating institute.

## Patient consent

waived

